# Longitudinal dynamics of the neutralizing antibody response to SARS-CoV-2 infection

**DOI:** 10.1101/2020.07.14.20151159

**Authors:** Kai Wang, Quan-Xin Long, Hai-Jun Deng, Jie Hu, Qing-Zhu Gao, Gui-Ji Zhang, Chang-Long He, Lu-Yi Huang, Jie-Li Hu, Juan Chen, Ni Tang, Ai-Long Huang

## Abstract

**Background:** Coronavirus disease 2019 (COVID-19) is a global pandemic with no licensed vaccine or specific antiviral agents for therapy. Little is known about the longitudinal dynamics of SARS-CoV-2-specific neutralizing antibodies (NAbs) in COVID-19 patients.

**Methods:** Blood samples (n=173) were collected from 30 COVID-19 patients over a 3-month period after symptom onset and analyzed for SARS-CoV-2-specific NAbs, using the lentiviral pseudotype assay, coincident with the levels of IgG and proinflammatory cytokines.

**Results:** SARS-CoV-2-specific NAb titers were low for the first 7-10 d after symtom onset and increased after 2-3 weeks. The median peak time for NAbs was 33 d (IQR 24-59 d) after symptom onset. NAb titers in 93.3% (28/30) of the patients declined gradually over the 3-month study period, with a median decrease of 34.8% (IQR 19.6-42.4%). NAb titers increased over time in parallel with the rise in IgG antibody levels, correlating well at week 3 (r = 0.41, p < 0.05). The NAb titers also demonstrated a significant positive correlation with levels of plasma proinflammatory cytokines, including SCF, TRAIL, and M-CSF.

**Conclusions:** These data provide useful information regarding dynamic changes in NAbs in COVID-19 patients during the acute and convalescent phases.

## Introduction

Coronavirus disease 2019 (COVID-2019) is a novel respiratory disease that is caused by severe acute respiratory syndrome coronavirus 2 (SARS-CoV-2). Since the outbreak of SARS-CoV-2 last year, it has spread rapidly and caused a global pandemic.^1^ As of July 3, 2020, over 10 million people worldwide have been reportedly infected and more than 512,000 individuals have died of COVID-19.^2^ Currently, considerable progress is being made to understand SARS-CoV-2 pathogenesis, epidemiology, antiviral drug development, and vaccine design. However, no licensed specific antiviral drugs or prophylactic vaccines are available. Developing effective viral inhibitors and antibody-based therapeutics to prevent or treat COVID-19 infection is a high global priority.

The SARS-CoV-2 RNA genome encodes 29 structural and non-structural proteins, including spike (S), envelope (E), membrane (M), and nucleocapsid (N) proteins, and the ORF1a/b polyprotein.^3^ The S glycoprotein is responsible for SARS-CoV-2 attachment and entry into target host cells via its binding to the angiotensin-converting enzyme 2 (ACE-2) receptor.^4^ Virus-specific neutralizing antibodies (NAbs) play a key role in reducing viral replication and increasing viral clearance.^5,6^ NAbs act against the receptor-binding domain (RBD) of the SARS-CoV-2 S protein, effectively blocking viral entry. Thus, serological testing, especially to detect NAbs, is essential in determining the onset of the serological immune response, evaluating the potential capacity of the host body for viral clearance, and identifying donors for passive antibody therapy trials.

In COVID-19 patients, NAbs can be detected within 2 weeks of symptom onset.^7,8^ The serological antibody response continues for at least 3 weeks and, in some cases, substantially longer.^9,10^ However, the dynamics and roles of SARS-CoV-2-specific NAbs and their correlation with antibody responses have not been explored in COVID-19 patients more than two months after symptom onset.

In this study, we first analyzed the 3-month longitudinal dynamics of in vitro NAb titers in 30 recovered COVID-19 patients. Second, we evaluated the correlation between the dynamics of NAb titers and serological IgG levels, as well as inflammatory cytokine levels. Our study may provide useful information regarding dynamic changes in NAbs in COVID-19 patients during the acute and convalescent phases and aid in the development of vaccines against SARS-CoV-2.

## Materials and methods

### Clinical characteristics

A total of 30 COVID-19 patients who had recovered and were discharged from the Yongchuan Hospital of Chongqing Medical University were included in our cohort. On April 2nd and May 8th, 2020, two follow-up visits were conducted to measure and characterize the dynamic changes in virus-specific IgG and NAb titers.

### Ethical approval

The study was approved by the Ethics Commission of Chongqing Medical University (ref. no. 2020003). Written informed consent was waived by the Ethics Commission of the designated hospital for emerging infectious diseases.

### Plasmids

The codon-optimized gene encoding the SARS-CoV S protein (AAP13567.1) and SARS-CoV-2 S protein (QHD43416) with the 19 C-terminal amino acids deleted were synthesized by Sino Biological Inc (Beijing, China) and cloned into the the pCMV3 vector, respectively. The HIV-1 NL4-3 ΔEnv Vpr luciferase reporter vector (pNL4-3.Luc.R-E-), constructed by N. Landau,^11^ was provided by Cheguo Cai, Wuhan University (Wuhan, China). The vesicular stomatitis virus G (VSV-G)-expressing plasmid pMD2.G was provided by Prof. Ding Xue, Tsinghua University (Beijing, China).

### Cell lines

HEK293T cells were purchased from the American Type Culture Collection (ATCC, Manassas, VA, USA). Cells were maintained in Dulbecco’s modified Eagle’s medium (DMEM; Hyclone, Waltham, MA, USA) supplemented with 10% fetal bovine serum (Gibco, Rockville, MD, USA), 100 mg/mL streptomycin, and 100 U/mL of penicillin at 37 °C in 5% CO_2_. HEK293T cells transfected with human ACE2 (293T-ACE2) were cultured under the same conditions, with the addition of G418 (0·5 mg/mL) to the medium.

### Production and titration of SARS-CoV-2 S pseudovirus

The SARS-CoV and SARS-CoV-2 pseudoviruses were generated as previously described, with some modifications.^12^ Briefly, HEK293T cells (5 × 10^6^) were co-transfected with pNL4-3.Luc.R-E- and recombinant SARS-CoV S or SARS-CoV-2 S plasmid using the Lipofectamine 3000 transfection reagent (Invitrogen, Rockville, MD), according to the manufacturer’s instructions. The cells were transferred to fresh DMEM 12 h later. The supernatant containing pseudovirions were harvested 48 h after transfection and passed through a 0·45-μm filter. To construct the VSV-G pseudovirus, pMD2.G was co-transfected with the pNL4-3.Luc.R-E-plasmid. Viral titers (RNA copy number, copies/mL) were determined by real-time RT-qPCR using primers targeting the long terminal repeat (LTR) region.^13^

### Neutralization assays

The 293T-ACE2 cells (2 × 10^4^ cells/well) were seeded in 96-well plates. For the neutralization assay, 50 μL of pseudovirus (3·8 × 10^4^ copies) was incubated with serial dilutions of serum samples from patients and human control serum as a negative control for 1 h at 37 °C and then added to the 96-well 293T-ACE2 plates. After 12 h of infection, fresh culture was added to each well. After 72 h post-infection, the 293T-ACE2 cells were lysed with 30 μL lysis buffer (Promega, Madison, WI, USA) to measure pseudoviral transduction. Relative luminescence units of Luc activity were determined using the Luciferase Assay Kit (Promega). Titers of NAbs were calculated as the 50% inhibitory dose (ID_50_).

### Statistical analysis

Continuous variables were expressed as median (inter-quartile range, IQR) and categorical variables were expressed as number (percentage, %). Comparisons between two groups were performed using the Mann-Whitney U test or Fisher’s exact test. A two-sided α of <0·05 was considered statistically significant. Statistical analyses were performed using *R* software, v3.6.0. Two-tailed Pearson correlation test was used to calculate the correlation coefficient of NAb to IgG levels or cytokines.

## Results

### Clinical characteristics

Of the total 30 patients in the cohort, 60·0% (18/30) were female, and 10·0% (3/30) were categorized as severe based on the COVID-19 Treatment guidelines (National Health Commission of the People’s Republic of China) (Table 1). The median length of the hospital stay was 22 d (IQR 15–26).

**Table 1.** Clinical characteristics of 30 patients enrolled in this study.

| **Characteristics** | **Patients (n = 30)** |
| --- | --- |
| **Sex (Male/Female)** |  |
| Male | 12 (40.0%) |
| Female | 18 (60.0%) |
| **Comorbidities** |  |
| Hypertension | 9 (30.0%) |
| Cardiovascular disease | 2 (6.7%) |
| Diabetes | 2 (6.7%) |
| COPD | 1 (3.3%) |
| Chronic kidney disease | 1 (3.3%) |
| Chronic liver disease | 2 (6.7%) |
| Any | 13 (43.3%) |
| **Signs and symptoms** |  |
| Fever | 11 (36.7%) |
| Fatigue | 2 (6.7%) |
| Dry cough | 10 (33.3%) |
| Inappetence | 2 (6.7%) |
| Myalgia | 3 (10.0%) |
| Dyspnea | 5 (16.7%) |
| Expectoration | 7 (23.3%) |
| Pharyngalgia | 5 (16.7%) |
| Diarrhea | 1 (3.3%) |
| Nausea | 1 (3.3%) |
| Dizziness | 2 (6.7%) |
| Vomiting | 1 (3.3%) |
| Chills | 3 (10.0%) |
| Rhinorrhea | 1 (3.3%) |
| Chest stuffiness | 1 (3.3%) |

### Specific serum response to SARS-CoV-2 pseudovirus infection

Sera from five representative COVID-19 convalescent patients, collected during the fourth week following symptom onset, were analyzed for their NAb titers against SARS-CoV-2 pseudovirus infection of 293T-ACE2 cells. All five serum samples demonstrated neutralizing activity against SARS-CoV-2 pseudovirus infection, with the ID_50_ value of 173.5, 267.9, 532.7, 730.4 and 1114, while the control serum from healthy individuals showed no neutralizing activity (Figure 1A and B). Furthermore, 30 plasma samples with strong SARS-CoV-2 neutralizing activity were evaluated for neutralization of VSV-G, SARS-CoV, and SARS-CoV-2 pseudoviruses. No detectable neutralization was found against the SARS-CoV and VSV-G pseudovirus control (Figure 1C). These results suggest that SARS-CoV-2 infection could not stimulate strong cross-neutralizing antibodies against SARS-CoV.

**Figure. 1.**
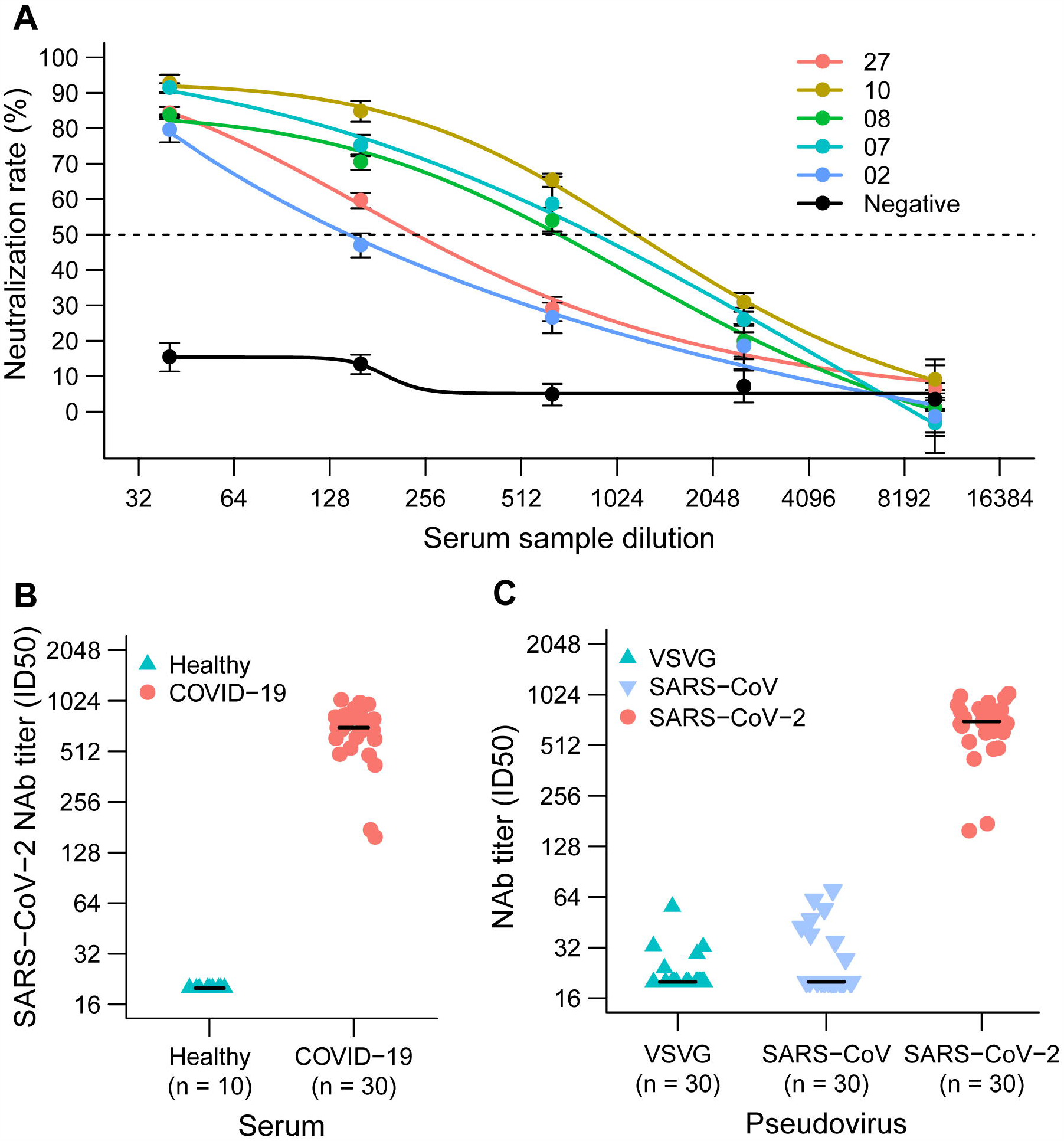
Analysis of the plasma response to SARS-CoV-2 infection. (A) Sera from five convalescent COVID-19 patients neutralized the SARS-CoV-2 pseudovirus. A serum sample from a healthy individual served as the negative control. The assay was performed in triplicate, and the median percentage of neutralization is shown. (B) SARS-CoV-2 neutralizing antibody (NAb) titers of 20 plasma samples from COVID-19 convalescent patients and ten plasma samples from healthy donors. (C) Neutralizing antibody titers against VSV, SARS-CoV, and SARS-CoV-2 pseudovirus in the sera from 30 convalescent COVID-19 patients. Student’s t-test, *P < 0.05, **P < 0.01, ***P < 0.001.

### Dynamic changes in neutralizing antibodies against SARS-CoV-2

We analyzed the longitudinal dynamics of virus-specific IgG and NAb levels in 30 patients who were positive for SARS-CoV-2 using real-time RT-qPCR. Sequential serum samples were collected from patients in the acute phase (5–6 samples) and the convalescent phase (two follow-up points: 60 d [54–63 d] and 96 d [90–99 d] after symptom onset). SARS-CoV-2-specific NAb titers were low before day 7–10 and increased at week 2–3 after symptom onset. The highest NAb levels were detected 3 months after symptom onset in 28 of 30 (93.3%) COVID-19 patients (Figure 2A). The median time for peak NAb levels was 33 d (IQR 24–59 d) after symptom onset, and the NAb levels plateaued in 60% (18/30) of the COVID-19 patients during hospitalization (Figure 2B). The peak NAb levels varied among the patients; 6·7%, 73·3%, and 20% patients showed low (ID_50_ < 500), medium-low (ID_50_ 500–999), and medium-high (ID_50_ 1000–2500) NAb titers, respectively (Figure 2C). There was no statistical difference among peak NAb titers that occurred during hospitalization and convalescence (Figure 2D).

**Figure 2.**
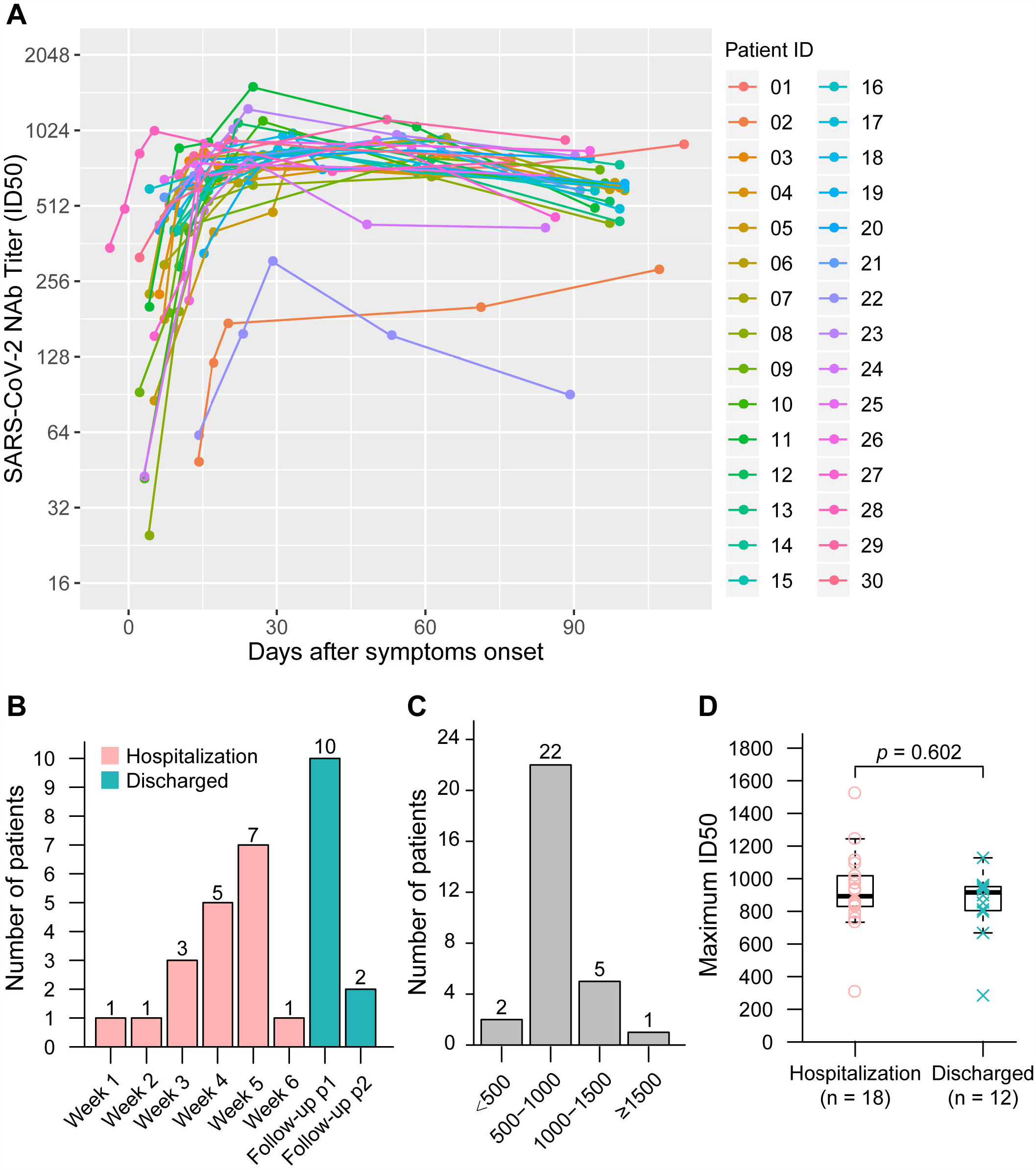
Dynamic changes in neutralizing antibodies against SARS-CoV-2. **A.** Kinetics of SARS-CoV-2 neutralizing antibody (NAb) levels in 30 COVID-19 patients. B. Number of patients experiencing peak NAb levels during hospitalization or after discharge. C. Peak NAb levels in the patients. D. Comparison of peak NAb levels between COVID-19 patients experiencing peak NAb levels during hospitalization and COVID-19 patients experiencing peak NAb levels after discharge.

The duration and maintenance of peak of NAb levels in COVID-19 patients is of great concern. Thus, we compared NAb levels between the peak time point and the final follow-up time point. A decline in NAb levels was observed in 93·3% (28/30) of SARS-CoV-2 infected patients, with a median decrease of 34·8% (IQR 19.6–42.4%) (Figure 3A). Patients were also grouped according to their rate of decrease in NAb levels; more than 20% of the patients showed a >70% decrease in NAb levels during this time period (21/30) (Figure 3B).

**Figure 3.**
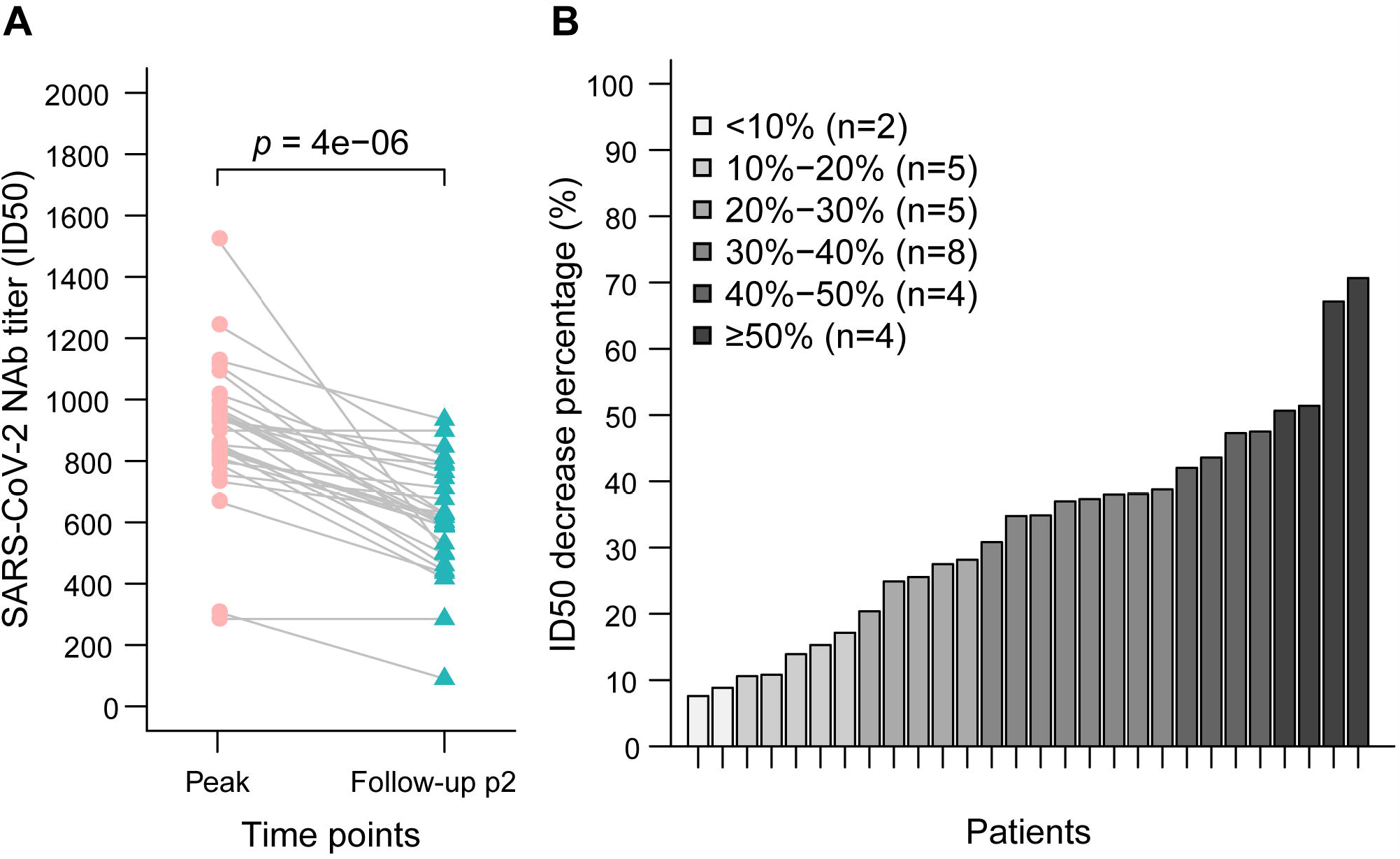
Decrease in neutralizing antibody levels in COVID-19 patients. A. Comparison of neutralizing antibody (NAb) levels between the highest time point and the final follow-up time point. B. Percentage decrease in neutralizing antibodies in COVID-19 patients.

### Correlation between dynamics of NAbs and IgG levels in COVID-19 patients

The kinetic levels of NAbs and virus-specific IgG over time in COVID-19 patients are still unknown. To address this, we first determined the relationship between aligned the NAb levels and virus-specific IgG levels in individual patients (Figure 4A and Supplementary Figure 1); similar dynamic changes were observed for the NAbs and virus-specific IgG levels in some patients. Furthermore, to determine if there was a statistical correlation between NAb levels and virus-specific IgG levels in COVID-19 patients, serum samples were grouped by time (weeks) after symptom onset. A statistically significant positive correlation was only observed in samples obtained 3 weeks after symptom onset (p = 0·027, r = 0·410) (Figure 4B).

**Figure 4.**
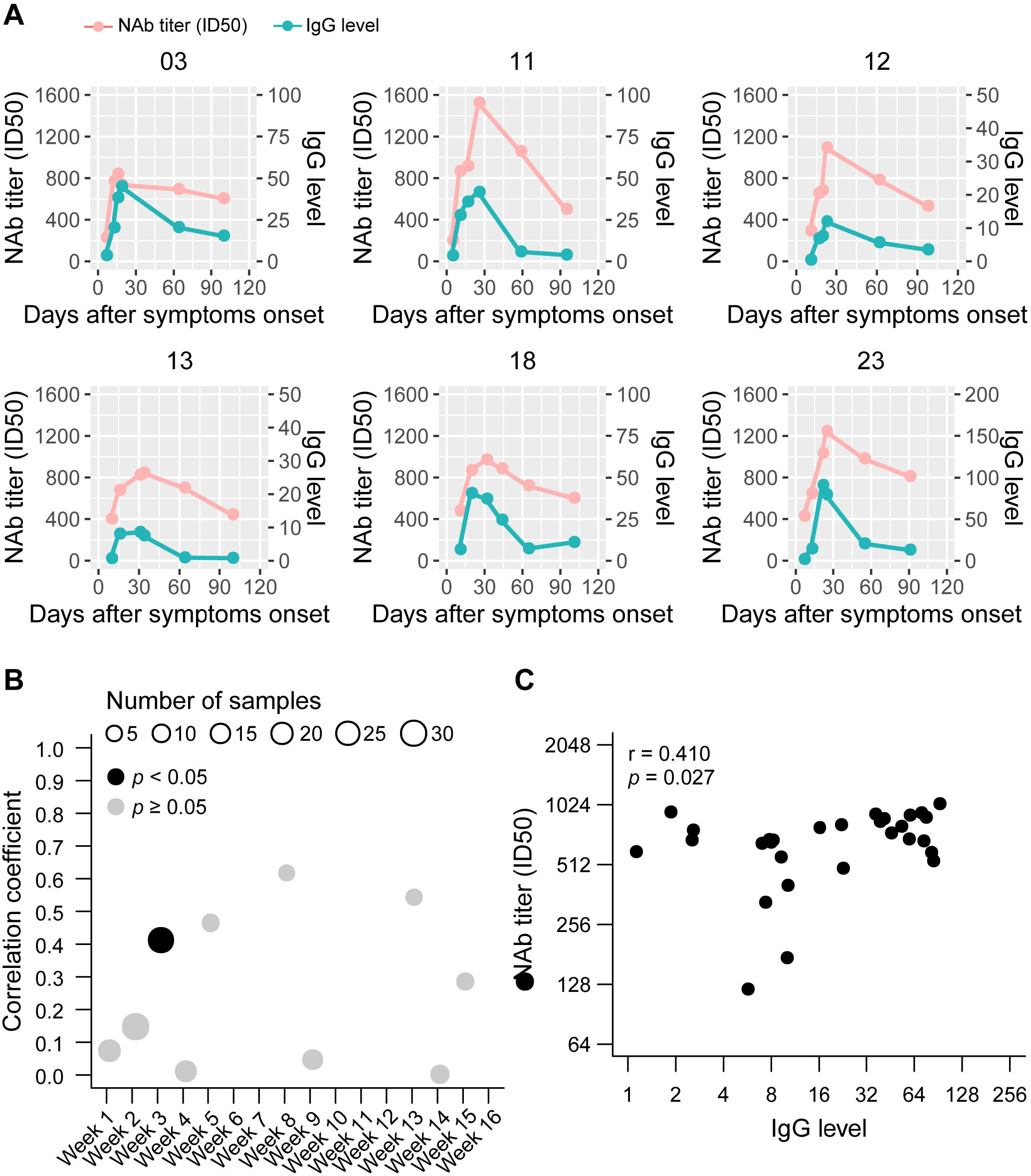
Correlation between the dynamics of neutralizing antibody and virus-specific IgG levels. A. Kinetics of neutralizing antibody (NAb) and IgG levels in six patients. Plasma samples were collected at different time-points after symptom onset. B. 152 serum samples were grouped by time of collection after symptom onset; correlations were analyzed between neutralizing antibody levels and IgG levels in each group. C. Correlations between neutralizing antibody levels and IgG levels from serum samples collected 3 weeks after symptom onset.

### Roles of cytokines in B cell function or antibody production

We analyzed the correlation between cytokine and chemokine levels and NAb levels in COVID-19 patients during the acute phase. Interestingly, we observed that NAb levels were positively correlated with stem cell factor (SCF) (r = 0·616, p = 0·001), TNF-related apoptosis-inducing ligand (TRAIL) (r = 0·514, p = 0·008), and macrophage colony-stimulating factor (M-CSF) (r = 0·454, p = 0·017) levels (Figure 5).

**Figure 5.**
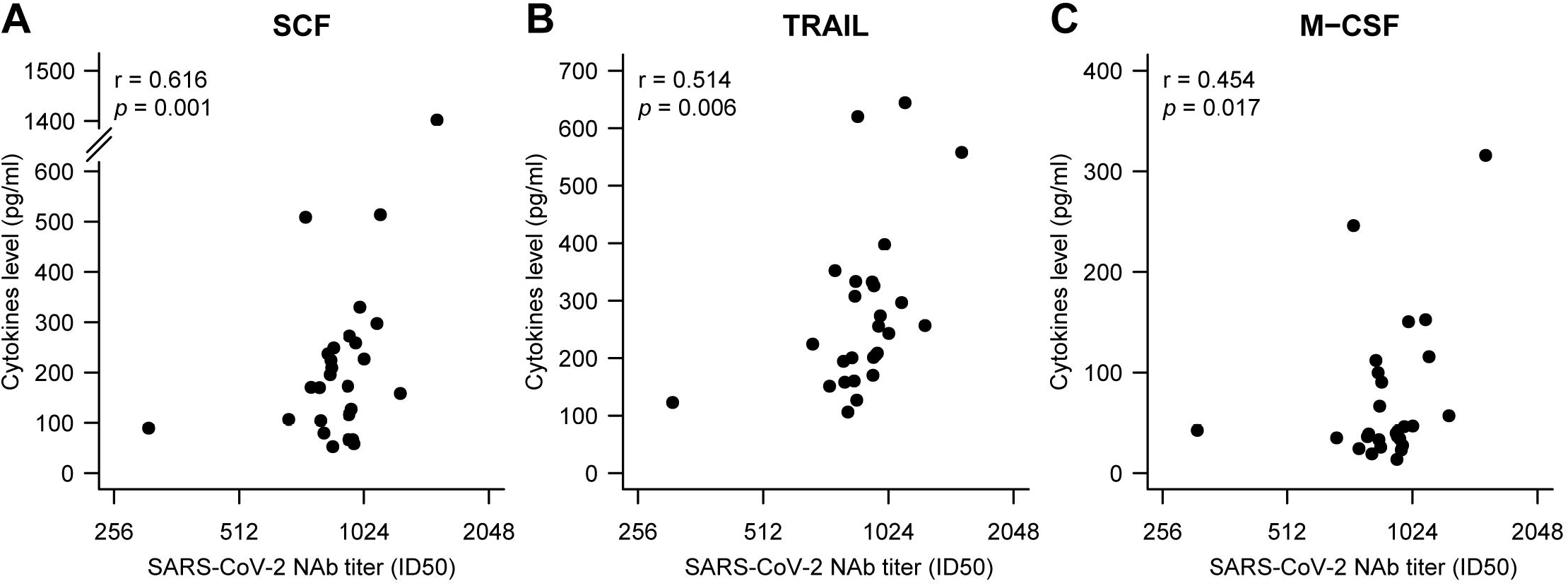
Correlation between peak neutralizing antibody levels and cytokines in sera. Serum samples with the highest neutralizing antibody (NAb) levels for all individuals were collected during hospitalization. Cytokine or levels were measured using the Bio-Plex Human Cytokine Screening Panel (48-Plex no. 12007283, Bio-Rad) on a Luminex 200 (Luminex Multiplexing Instrument, Merck Millipore), following the manufacturer’s instructions. Pearson’s correlation was used to analyze differences between NAb levels and cytokine levels.

## Discussion

Virus-specific NAbs have been considered an important determinant for viral clearance. The pseudovirus-based assay is suitable for the high-throughput screening of SARS-CoV-2 NAbs in plasma donors without the requirement of BSL-3 laboratories. The assay has been widely used for evaluating NAbs in highly pathogenic viruses, such as Ebola, SARS-CoV, MERS-CoV, and highly pathogenic influenza viruses.^14^ Herein, we described the dynamics of SARS-CoV-2-specific NAbs generated during both the acute and convalescent phases of SARS-CoV-2 infection using a pseudovirus-based neutralization assay. We found that SARS-CoV-2-specific NAb titers were low before day 7–10, peaked at approximately day 33 after symptom onset, and then gradually declined over a 3-month period. Meanwhile, SARS-CoV-2-specific NAbs were detected concurrently with and positively correlated with IgG antibodies in our cohort, indicating that the NAb response may play an important role in viral clearance.

Our understanding of the duration and nature of protective immunity to SARS-CoV-2 is currently very limited. The kinetics of antibody-mediated immunity to SARS-CoV-2 infection and how long this immunity lasts are unknown. Our data suggest that NAb titers in patients were variable, and the protective humoral immune response to SARS-CoV-2 may abate over time, which is in accordance with findings in patients infected with other human coronaviruses, such as HCoV-229E.^15,16^ The short-term humoral immune response in COVID-19 patients is also highly consistent with that observed in patients infected with SARS-CoV and MERS-CoV,^17,18^ who show a rapid decrease in virus-specific antibody titers within 3–4 months.

Among the 30 recovered patients in our study, two patients showed very low NAb titers during the acute phase and 3-month follow-up, indicating that other immune responses, involving T cells and inflammatory cytokines may have contributed to viral clearance. These data suggest that the antibody titers may diminish with time or some recovered patients may not produce a high-titer response during SARS-CoV-2 infection.

Recently, in a rhesus macaques model, SARS-CoV-2 infection evoked a robust protective immune response when the animals were re-exposed to SARS-CoV-2 one month after the initial viral infection.^19^ However, natural infection and volunteer challenge studies hint that coronavirus infections, including those with HCoV-229E and HCoV-OE43, cannot induce stable protective immunity; thus, reinfection occurs frequently. Moreover, a SARS-CoV antigen-specific memory B cell response was not detectable in recovered SARS patients at 6-years after disease onset, whereas SARS-CoV-specific memory T cells persisted in recovered SARS patients.^20,21^ Although the role of memory T cells in the protective immune response to SARS-CoV-2 needs further evaluation, a robust T cell response is required for viral clearance.

We also described here, the dynamic correlation between SARS-CoV-2-specific NAbs and serological total IgG levels. NAb titers appeared concomitantly and correlated moderately with IgG levels at week 3 after symptom onset, which is consistent with other reports regarding COVID-19 recovered patients^10,22^. The antigen epitope used for IgG detection in our study contained the nucleoprotein peptide, as well as the RBD domain of the spike protein, which partially explains the discrepancy in NAb titers and IgG levels at weeks 4, 9, and 14 after symptom onset. The nucleoprotein is the most abundant protein in the SARS-CoV-2 viral particle and possesses the strongest immunogenicity. The binding antibodies detected by the total IgG assay may also be involved in viral clearance through antibody-dependent cytotoxicity, Therefore, the roles of binding antibodies and NAbs in disease progression need further evaluation.

Currently, adaptive immunotherapy using convalescent plasma (CP) from recovered COVID-19 patients is being employed as a potential therapeutic approach to confer antiviral protection.^23^ Several preliminary clinical trials have proven its effectiveness in treating SARS-CoV-2.^6,24^ The efficacy of CP transfusion is attributed to the neutralizing effect of antibodies; thus, the NAb titer is the major determinant for CP therapy. Monitoring NAb levels and their duration will provide valuable data for evaluating the effectiveness of CP therapy. In our study, the levels of NAbs declined gradually over the 3-month follow-up period, with a median decrease of 34.8%. Thus, CP samples with high titers of NAbs from patients in the early stage of convalescence will be more suitable for clinical use.

There are some limitations to this study, which should be addressed. Due to the small sample size, we could not find any correlation between the dynamics of NAb titers and clinical characteristics contributing to different clinical outcomes. Serological blood samples were collected up to 3 months after symptom onset; data collected over longer follow-up times should be obtained to demonstrate the duration of humoral immunity after SARS-CoV-2 infection. The lack of data to determine an anamnestic immune response, such as tests for SARS-CoV-2-specific memory B cells, memory T cells, and specific cytokine-dependent memory cells, hampered the evaluation of the immune response, especially protective immunity against viral reinfection. These are major issues that should be investigated in future studies.

In summary, we determined the dynamics of NAb titers within 3 months after symptom onset in 30 SARS-CoV-2-infected patients and found a positive correlation between NAb titers and IgG antibodies. Our work provides valuable insight into the humoral immunity against SARS-CoV-2 infection. We also described a pseudotype system for measuring NAb titers, which could be expanded to antiviral drug screening and vaccine development.

## Data Availability

The data supporting the findings of this study are available from the authors upon request.

## NOTES

## Acknowledgments

We would like to thank Prof. Cheguo Cai (Wuhan University, Wuhan, China) for providing the pNL4-3.Luc.R-E-plasmid.

